# PSYCHOLOGICAL FACTORS PREDICTING OUTCOMES OF CONSERVATIVE TREATMENT IN FROZEN SHOULDER PATIENTS (PSY-FS)

**DOI:** 10.1101/2022.11.12.22282202

**Authors:** Fabrizio Brindisino, E Silvestri, A Fioretti, G Giovannico, G Di Giacomo, D Feller, A Chiarotto

## Abstract

Frozen shoulder contracture syndrome (FSCS) is characterized by underhand onset, severe shoulder daily and night pain, active and passive range of motion (ROM) limitation, disturbing sleep, and shoulder-related disability.

FSCS has a cryptogenetics etiology and is more prevalent in people with diabetes, autoimmune and thyroid disease, with higher prevalence in the age range between 50 and 60. Notably no deal is about higher incidence in people with physically low activity and female sex. Moreover, although some patients report complete symptom recovery, others report residual motion impairments and pain.

Research on prognostic factors was applied on FSCS, and Eljabu et colleagues (2016) stated that diabetes, comorbidities, bilateral presentation and onset higher pain and disability are negative prognostic factors that could direct patient to early surgery; however, little is known about the prognostic influence of psychological factors in FSCS patients. On the other hand, in other shoulder pathologies, the presence of psychological factors is well documented, and evidence confirms that some of these features could represent prognostic factors that impact the prognosis.

A recent systematic review reported that psychological factors were associated with increased pain perception and decreased function and quality of life at baseline in patients with FSCS, and pain-beliefs seem to be associated with a worst perception of arm function; however, little is known about the prognostic value of such factors in FSCS recovery.

Knowing about the presence and the role of all types of prognostic factors is important because they can aid treatment and lifestyle decisions, improving individual risk prediction, providing novel targets for new treatments, and enhancing collaboration between different professionals.

This study aims to determine if pain, function, disability, quality of life, ROM and time for recovery were influenced by psychological factors in FSCS patients.

---

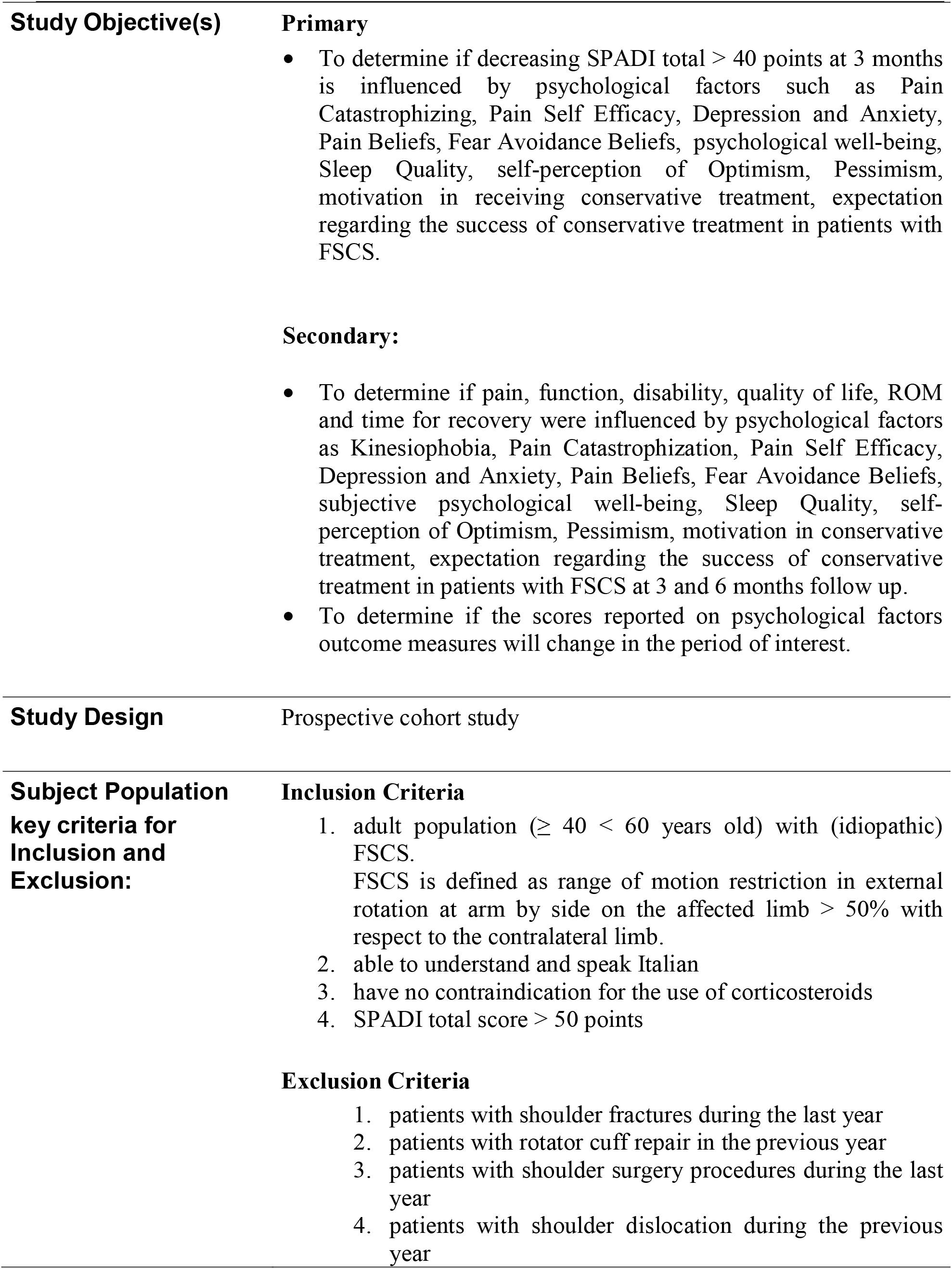
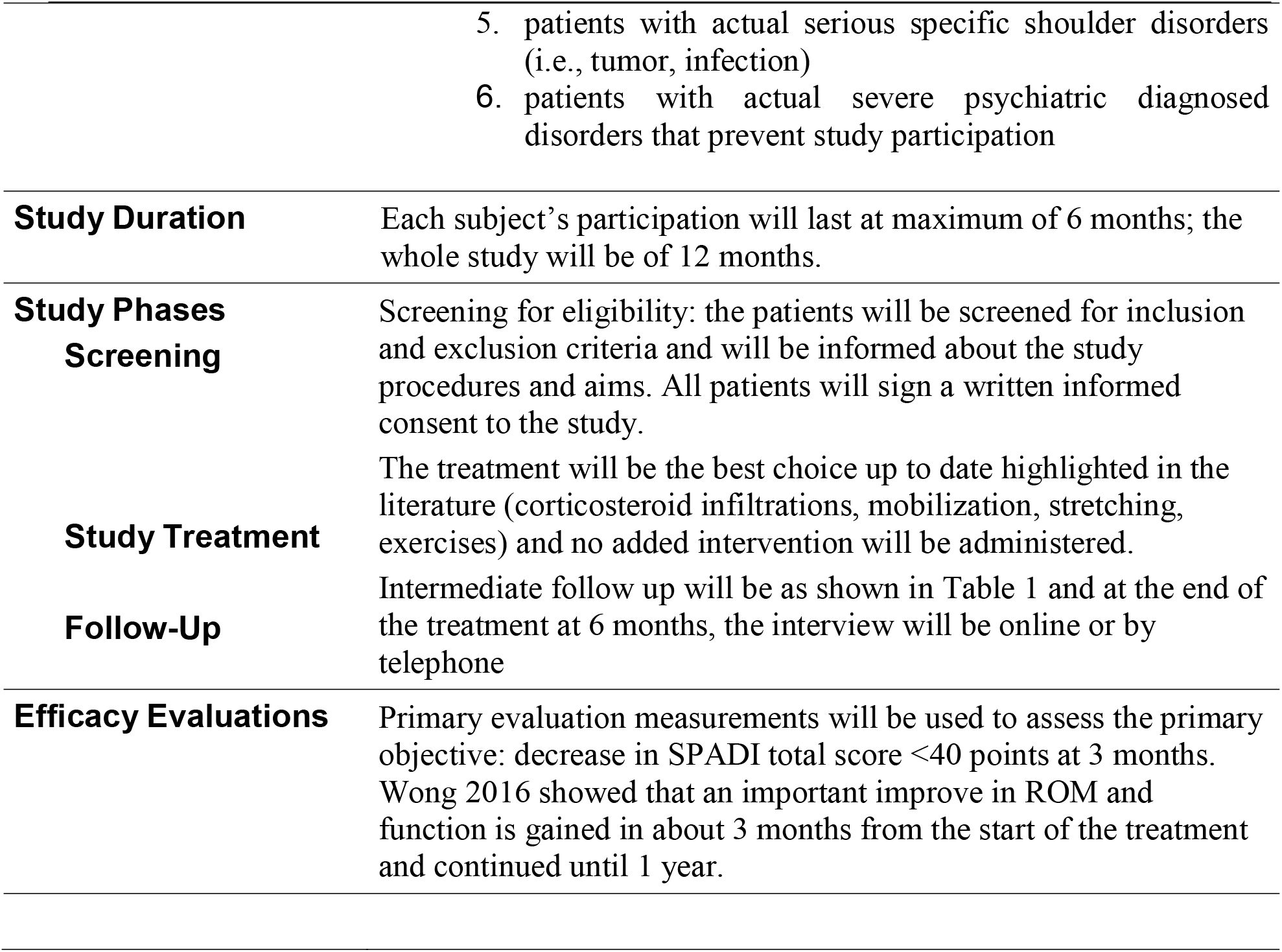

## Section A- DEMOGRAPHIC INFORMATION REQUESTED

Age Gender

Work activity

predominantly manual heavy workers

Education

Physical activity

Physical activity with affected shoulder

Dominant arm

Affected arm BMI

FS in the past (same shoulder)

FS in the past (contralateral shoulder)

Comorbidities (present)

Comorbidities (past)

Drugs

### Pain and Stiffesss assessment

Pain onset

Daily pain

Nightly pain

Mean pain in the previous week

Pain Onset (time)

Stiffness onset (time)

Time between symptoms onset and first medical assessment

Time between symptoms onset and FS diagnosis

COVID-19 infection

COVID-19 vaccination(s)

## Section B- PHYSICAL EXAMINATION PROCEDURES

Inclinometric evaluaton of ROM assessment in:

gleno-humeral ER 0° ABD

gleno-humeral ABD

gleno-humeral FLEX sagittal plane

gleno-humeral retroposition (maximum vertebrae level reached/cm from C7 spinous process).

IR in “sleeper stretch” position

## Section C- PROMs ADMINISTRATED (outcome)

NRS for PAIN (at rest, activity and night)

NRS for PERCEIVED STIFFNESS

SPADI Pain/SPADI disability/SPADI total (for pain and disability)

EURO-QoL 5D 5L

DASH (for Manual Functioning, Shoulder Range of Motion, and Symptoms and Consequences subscales)

## Section D- PROMs ADMINISTRATED (exposure)

NRS-for fear of pain

Central Sensitization Inventory (CSI)

Pain Catastrophizing Scale (PCS)

Pain Self Efficacy Questionnaire (PSEQ)

Beck Depression Inventory 2 (BDI II)

STAI y1; STAI y2

Pain Beliefs and Perception Inventory (PBAPI)

Fear Avoidance Beliefs Questionnaire (FABQ)

subjective psychological well-being (WHO-5)

Pittsburgh Sleep Quality Index (PSQI)

Insomnia Severity Index (ISI)

NRS for self-perception of Optimism

NRS for self-perception of Pessimism

NRS for self-perception of motivation in conservative treatment

NRS for self-perception of expectation regarding the success of conservative treatment

NRS for satisfaction and level of satisfaction for conservative approach

## PROMs DESCRIPTION

### Section C

#### Pain intensity (numeric pain rating scale – NRS-pain) (Hawker et al., 2011)

The NPRS is an 11-point numeric rating scale, to indicate the shoulder pain intensity, with 0 = no pain at all and 10 = worst imaginable pain. Participants were asked to score their average shoulder pain intensity during the present week at rest, at night and while performing activities of daily living.

#### Perceived stiffness (numeric stiffness rating scale - NRS-stiffness) (Bacci et al., 2017)

An 11-point numeric rating scale was used to assess the perceived shoulder stiffness during movement, with 0 = no feelings of shoulder stiffness and 10 = feeling of highest imaginable shoulder stiffness. Participants were asked to score their average perceived shoulder stiffness, thinking of their arm movement during the present week.

#### Disabilities of the Arm, Shoulder, and Hand Questionnaire (DASH) (Padua et al, 2003)

The DASH is composed of 30 questions measuring physical function, social function, and symptoms in patients with musculoskeletal disorders of the upper limb. Each item is rated with a 5-points Likert scale. As the DASH Italian version (Padua et al., 2003) demonstrated a three-factor structure (Franchignoni et al., 2010), three different scores will be calculated: items 1–5, 7–11, 16–18, 20, and 21 composed the Manual Functioning subscale, items 6, 12–15, and 19 composed the Shoulder Range of Motion subscale, and items 22–30 composed the Symptoms and Consequences subscale. The total score of each subscale was computed by adding the scores attributed to each item.

#### Shoulder Pain and Disability Index – SPADI- (Giannotta et al., ahead of print)

The SPADI was developed to measure the pain and disability associated with shoulder pathology. The SPADI is a self-administered index consisting of 13 items divided into two subscales: pain and disability; a 5-item subscale that measures pain and an 8-item subscale that measures disability

##### EURO-QoL 5D 5L *(2009 EuroQol Group EQ-5D)*

The 5-level EQ-5D version (EQ-5D-5L) was introduced by the EuroQol Group in 2009 to improve the instrument’s sensitivity and to reduce ceiling effects, as compared to the EQ-5D-3L. The EQ-5D-5L essentially consists of 2 pages: the EQ-5D descriptive system and the EQ visual analogue scale (EQ VAS). The descriptive system comprises five dimensions: mobility, self-care, usual activities, pain/discomfort and anxiety/depression. Each dimension has 5 levels: no problems, slight problems, moderate problems, severe problems and extreme problems. The patient is asked to indicate his/her health state by ticking the box next to the most appropriate statement in each of the five dimensions.

The EQ VAS records the patient’s self-rated health on a vertical VAS, where the endpoints are labelled ‘The best health you can imagine’ and ‘The worst health you can imagine’. The VAS can be used as a quantitative measure of health outcome that reflect the patient’s own judgement.

### Section D

#### Pain Catastrophizing (Pain Catastrophizing Scale - PCS) (Monticone, 2012)

The PCS is a 13-item scale to quantify negative thoughts that may be experienced in the presence of pain. Each question is scored on a 5-point Likert scale (from “not at all” to “always”). The total score of the PCS ranges between 0 and 52 with a higher score indicating greater pain catastrophizing.

#### Self-Efficacy (Pain Self Efficacy Questionnaire – PSEQ) (Chiarotto, 2015)

The PSEQ is used to measure pain self-efficacy and consists of 10 items representing different daily activities (eg, I can do most of the household chores) or general aspects of life (eg, I can still accomplish most of my goals in life). For each item, the patient must rate how confident he or she feels to perform these activities, despite the presence of pain. Items are rated on a 7-point Likert scale ranging from 0 (not at all confident) to 6 (completely confident). The total score of the questionnaire can range from 0 to 60, with higher scores indicating higher pain self-efficacy.

#### Back Depression Inventory II (Sica, 2007)

Beck Depression Inventory II Edition (BDI-2) is a 21-item multiple-choice self-report inventory for assessing affective-somatic (AS) and cognitive dimensions (C) of depression severity. The total score ranges from 0 to 63; in particular, a score from 10 to 18 indicates a mild to moderate depression, a score from 19 to 29 indicates a moderate to severe depression, and a score higher than 30 is indicative of a severe depression level.

#### STAI Y1Y2 (Ilardi, 2021)

State-Trait Anxiety Inventory (STAI Form Y1 and Y2) is a 40-item multiple-choice self-report inventory divided into two sub-tests, each having 20 items (for both state- and trait-anxiety) aimed at assessing and quantifying anxiety disorder in adults. All items are rated on a 4-point Likert scale. The recommended clinical cut-off is >46.

#### Pain Beliefs and Perceptions Inventory (PBAPI) (Monticone, 2013)

This is a 16-item questionnaire and patients rate their beliefs using a 4-point Likert scale ranging from -2 (total disagreement) to -2 (total agreement); item Nos. 3, 9, 12 and 15 are reverse scored. For each subscale, the scores of the responses to the items that are answered are added and divided by the number of items answered; higher scores indicate greater endorsement of the beliefs.

#### Fear Avoidance Beliefs Questionnaire (FABQ) (Monticone, 2012)

The FABQ-I has a 2-factor, 12-item structure and is reliable, valid, and sensitive to change, with the results replicating those of the other existing versions. It can therefore be recommended for clinical and research purposes because it is expected to improve the cognitive-behavioral assessment. The 2 factors were called FABQ-Work (items 6, 7, 9, 10, 11, 12, and 15) and

FABQ-Physical activity (items 1, 2, 3, 4, and 5). The subscores are 0-42 for FABQ-work, and 0-30 for FABQ-physical activity, with a higher value reflecting a higher degree of fear-avoidance beliefs. The MDC was 12, reflecting the smallest change in score that is likely to reflect a true change rather than a measurement error. The paper of Mintken et al, 2010, support the measurement of pain-related fear in shoulder pain using a modified FABQ.

#### WHO-5 (Psychiatric Research Unit, WHO Collaborating Center for Mental Health, Frederiksborg)

The 5-item World Health Organization Well-Being Index (WHO-5) is a short and generic global rating scale measuring subjective well-being. The respondent is asked to rate how well each of the 5 statements applies to him or her when considering the last 14 days. Each of the 5 items is scored from 5 (all the time) to 0 (none of the time). The raw score therefore theoretically ranges from 0 (absence of well-being) to 25 (maximal well-being). Because scales measuring health related quality of life are conventionally translated to a percentage scale from 0 (absent) to 100 (maximal), it is recommended to multiply the raw score by 4.

#### Pittsburgh Sleep Quality Index (PSQI) (Curcio, 2013)

The 19-item Pittsburgh Sleep Quality Index (PSQI) is probably the most used retrospective self-report questionnaire, that measures sleep quality over the previous month. Seven clinically derived domains of sleep difficulties (sleep quality, sleep latency, sleep duration, habitual sleep efficiency, sleep disturbances, use of sleeping medications, and daytime dysfunction) are assessed by the questionnaire. Taken together, these sleep domains are scored as a single factor of global sleep quality. Usually, a global score higher than 5 is considered as an indicator of relevant sleep disturbances in at least two components or of moderate difficulties in more than three components. More recently, an overlapping of some components has been observed and three distinct factors have been extracted: sleep efficiency (including sleep duration and habitual sleep efficiency), perceived sleep quality (including subjective sleep quality, sleep latency and use of sleeping medication), and daily disturbances (including sleep disturbances and daytime dysfunction)

#### Insomnia Severity Index (ISI) (Castronovo, 2016)

ISI is a seven items questionnaire that assess, during the previous 2 weeks: the severity of sleep onset (item 1a), the severity of sleep maintenance (item 1b), early morning awakenings (item 1c), satisfaction level with current sleep pattern (item 2), interference with daily living (item 3), noticeability of impairment due to the sleep difficulty (item 4), level of distress caused by the sleep problem (item 5). The total score ranges from 0 to 28, with higher score indicating greater insomnia severity. Each item is rated on a 5-point Likert scale of 0–4 (for items 1–3, 0 = no problem, 4 = very severe problem; for item 4, 0 = very satisfied, 4 = very dissatisfied; for items 5–7, 0 = not at all, 4 = very much). A cut-off value of 15 has been used as threshold for a clinically relevant insomnia. The total score of the questionnaire is divided as follows: 0–7, no significant insomnia; 8–14 subthreshold insomnia; 15–21, moderate insomnia and 22–28 severe insomnia.

#### Central sensitization Inventory (CSI) (Chiarotto et al., 2018)

The Central Sensitization Inventory (CSI) is a patient-reported instrument designed to identify patient’s symptoms related to central sensitization. It consists of two parts: part A provides a 0-100 total score for 25 items on current health symptoms with five response options ranging from ‘never’ (0) to ‘always’ (4); part B investigates if patients were previously diagnosed by a physician with one or more of seven different conditions. The CSI showed satisfactory test-retest reliability and internal consistency in patients with various chronic pain conditions and healthy participants.

## 1. Compliance Statement

This study will be reported in accordance to the REMARK reporting guideline.

## 2.

## 1. General Schema of Study Design

prospective cohort study

### 1.1 Study Duration, Enrollment and Number of Sites

#### 1.1.1 Duration of Study Participation

The study duration per subject will be up to 90 days, with follow-up divided as showed in Table 1.

**Table 1:** Schedule of Study Procedures.

| **Study Phase** | **Screening** | **Observation Study Visits** | **1 Follow-Up Visit (telephone/email survey)** |
| --- | --- | --- | --- |
| **Visit Number** | **1** | **2** | **6** |
| **Study Days** | **1** | **90** | **180** |
| Screening the patient for Inclusion/Exclusion Criteria | X |  |  |
| Informed Consent signed | X |  |  |
| Demographics/Medical History (see below for full information requested) | X |  |  |
| Physical Examination (see below for full information requested) | X | X |  |
| PROMs administration (see below for full PROMs administered) | X | X | X (PROMs for “exposure” only) |

## 2 STUDY PROCEDURES

1. individuation of probably enrolled subject
2. screening for inclusion exclusion criteria
3. willing to participate and informed consent signed
4. collection of demographic variables, administration of PROMs for exposition and outcome; assessing for ROM
5. Treatment
6. evaluations at follow up points

### 2.1 Screening Visit (*Baseline*)

⍰ Screening for inclusion and exclusion criteria
⍰ Informed Consent
⍰ Collection of demographic variables
⍰ Physical assessment
⍰ PROMs administration

### 2.2 Treatment Period

Patient will be treated following the rehabilitation procedures descripted by Venturin et al., 2021 *(Venturin D, Brindisino F, Ristori D, Rossi A, Vascellari A, Poser A. The use of corticosteroid/anesthetic injections in conjunction with physical therapy in the treatment of idiopathic frozen shoulder: a case series. JOSPT Cases 2021;1(4):248-265. doi:10*.*2519/josptcases*.*2021*.*9960)*

Intrarticular injection will be administered. In particular 3 intrarticular injections (one injection every two weeks, for six weeks) of 1 cc of Kenacort diluted in 4 cc of physiological water.

Injections will be eco-guided with posterior joint access. Injections will be performed by the same orthopaedist with 30 yers of experiences in shoulder surgery and injection treatment.

#### 2.2.1 Visit 2 (90 days)

⍰ Physical Examination (ROM)
⍰ PROMs administration

#### 2.2.2 Visit 3 (180 days - Follow Up questionnaire telephone/email survey)

⍰ PROMs administration
⍰ Assess possible adverse events
⍰ Evaluation of drug intake during the treatment (out of usual medication intake and corticosteroid injection)

## STATISTICAL ANALYSIS

The shape of the distribution of the variables related to the population will be evaluated through a graphical analysis.

The collected variables will be described using descriptive statistics. In particular, mean and standard deviation, median with the interquartile range (IQR), and relative frequency and percentages will be calculated for the variables with normal, non-normal and categorical distribution, respectively.

To describe the changes in clinical and functional variables between admission and discharge, graphical analyses will be used. Also, we will report descriptive statistics at the various follow-ups (Table 1).

Lost at follow-up and drop outs will be counted and evaluated. Missing data (referable to exposure and outcome variables) will be imputed through a “multiple imputation” procedure. We will impute a number of datasets equal to the number of subjects with at least one missing data. The reasons of drop out will be presented if the research team is aware of them. Patients who withdraw their informed consent during follow-up will not be included in the analyses. Baseline characteristics of patients lost to follow-up will be compared with those remaining in the study.

## PRIMARY ANALYSIS

All the variables reported in Table 2 will be evaluated as possible prognostic factors in a univariable logistic regression model, using the 40-point “loss” in the SPADI assessment as the dependent variable.

**Table 2:**
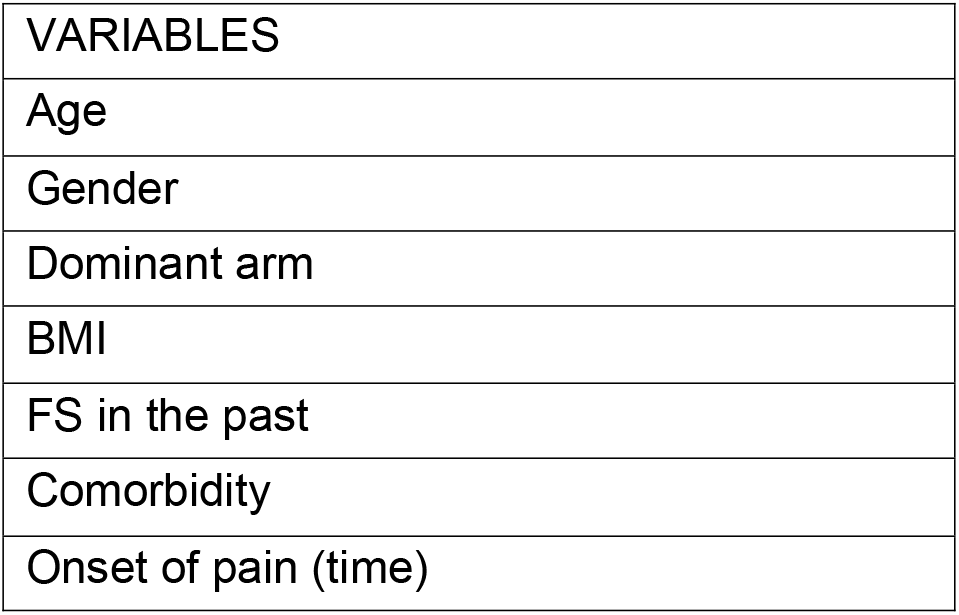
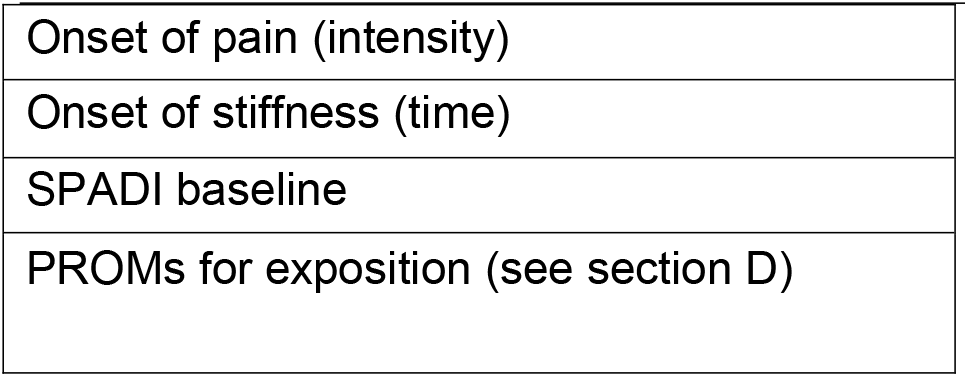
prognostic factors investigated/independent variables

| VARIABLES |
| --- |
| Age |
| Gender |
| Dominant arm |
| BMI |
| FS in the past |
| Comorbidity |
| Onset of pain (time) |
| Onset of pain (intensity) |
| Onset of stiffness (time) |
| SPADI baseline |
| PROMs for exposition (see section D) |

Following, we will perform a multivariable logistic regression model using the 40-point “loss” in the SPADI assessment (Sharma et al., 2017) as the dependent variable, while the independent variables will include all the known prognostic factors for FSCS (i.e.; intensity of pain and disability at the onset, comordibities, diabetes, and bilateral limb involvement, previous frozen shoulder, ER<0°, age < 60 y.o.).

The assumption of absence of multicollinearity will be evaluated through a correlation matrix (threshold value ρ> 0.9) and the Variance inflation factor (VIF) (threshold value> 10).

The possible presence of influencing values will be evaluated with the complex index “Cook distance” using 4/n as a threshold. All cases considered influencing to the estimates of the regression coefficients will be removed from the model estimation process by producing a model used for sensitivity analysis.

The linearity assumption between all continuous independent variables and the logit of the dependent variable will be checked visually using scatterplots and statistically with the Box - Tidwell test.

## SECONDARY ANALYSIS

For the secondary analysis, we will use the same approach as the primary analysis.

We will perform a linear regression for continuous dependent variables, while for time-to-event variables we will perform a survival analysis.

All statistical analyzes will be conducted with R.

### 2.3 Subject Completion/Withdrawal

Subjects may withdraw from the study at any time without prejudice to their care. They may also be discontinued from the study at the discretion of the Investigator for lack of adherence to study treatment or visit schedules, adverse events, or any other reason (to be documented). The Investigator may also withdraw subjects who violate the study plan, or to protect the subject for reasons of safety or administrative reasons. It will be documented whether each subject completes the clinical study. If the Investigator becomes aware of any serious, related adverse events after the subject completes or withdraws from the study, he will record it.

### 2.4 Clinical Adverse Events

Clinical adverse events (AEs) will be monitored throughout the study and assessed at 6 months.

### 2.5 Definition of an Adverse Event

An AE is any untoward medical occurrence in a subject who has received an intervention (drug, biological, or other intervention). The occurrence does not necessarily have a causal relationship with the treatment. An AE can therefore be any unfavorable or unintended sign (including an abnormal laboratory finding, for example), symptom, or disease temporally associated with using a medicinal product, whether considered related to the medicinal product.

All AEs (including serious AEs) will be noted in the study records and on the case report form with a complete description including the nature, date and time of onset, determination of non-serious versus serious, intensity (mild, moderate, severe), duration, causality, and outcome of the event.

## Data Availability

All data produced in the present study are available upon reasonable request to the authors

